# Clinical Validation and Machine Learning Optimization of MyCog: A Self-Administered Cognitive Screener for Primary Care Settings

**DOI:** 10.1101/2025.04.17.25325948

**Authors:** Stephanie Ruth Young, Yusuke Shono, Katherina Hauner, Elizabeth M. Dworak, Maxwell Mansolf, Laura Curtis, Julia Yoshino Benavente, Stephanie Batio, Richard C. Gershon, Michael S. Wolf, Cindy J. Nowinski

**Affiliations:** Department of Medical Social Sciences, Northwestern University Feinberg School of Medicine, Chicago, IL; School of Community and Global Health, Claremont Graduate University, Claremont, CA; Department of Neurology, Northwestern University Feinberg School of Medicine, Chicago, IL; Center for Applied Health Research on Aging, Northwestern University Feinberg School of Medicine, Chicago, IL

**Keywords:** Cognitive impairment, Alzheimer’s disease, primary care

## Abstract

**BACKGROUND:** Primary care presents an ideal opportunity for early detection of cognitive impairment, yet primary care clinics face barriers to cognitive screening. MyCog, an EHR-integrated tablet app that is self-administered during the rooming process of a primary care visit, streamlines the screening process to reduce barriers and encourage broader screening.

**METHODS:** We compared MyCog performance from 65 adults with diagnosed cognitive impairment to 80 cognitively normal adults, all aged 65+, recruited from clinical settings. We leveraged the consensus of five machine learning models (LASSO, Elastic Net, Random Forest, Bayesian Logistic Regression, and Gradient Boosting) to select consistently discriminative variables for the final detection algorithm. Performance was assessed at for two evidence-based thresholds (Youden’s J and Top Left) with ROC AUC, sensitivity, specificity, and accuracy as the primary metrics.

**RESULTS:** All five models showed strong diagnostic performance, with ROC AUC values ranging from 0.839 to 0.876. The consensus modeling approach consistently identified the MyCog Picture Sequence Memory (PSM) exact match score and the MyCog Dimensional Change Card Sort (DCCS) overall rate-correct score as predictors of cognitive impairment. The final logistic regression achieved a robust AUC of 0.890. Depending on the cut point selected, sensitivity ranged from 0.723−0.785 (95% CI: 0.547−0.877), specificity 0.825−0.912 (95% CI: 0.716−0.954), accuracy 0.807−0.828 (95% CI: 0.731−0.869).

**DISCUSSION:** MyCog provides practical, accurate cognitive screening for primary care. The sub-7-minute self-administered assessment eliminates staffing requirements and automates evaluation, addressing screening barriers to facilitate earlier detection and improve clinical outcomes. The algorithm’s robust performance and parsimony demonstrate clinical utility while maintaining diagnostic accuracy.

## Background

Cognitive impairment (CI) in older adulthood is a national and global health crisis,^1^ which may result from Alzheimer’s disease (AD) and AD-related dementias (ADRD)^2–5^, neurological disorders (e.g., Parkinson’s disease^6^, multiple sclerosis^7^), or reversible causes such as infections or medications^8^. Early detection of CI can help identify reversible causes, address symptoms and comorbidities, determine appropriate caregiver involvement, and help families plan for the future^9,10^. Primary care visits offer an important opportunity for early detection of CI in adults over age 65^11^, yet primary care practices face barriers to screening, such as lack of time, clinic resources^12^, and clinical training on CI management^13^.

Another underlying barrier to early detection efforts is the lack of standardized, minimally burdensome cognitive assessments that are both sensitive to early indicators of impairment and appropriate for use in resource-constrained primary care settings^14–17^. The most common traditional paper-and-pencil screeners, such as the Montreal Cognitive Assessment (MoCA)^18^, Mini-Cog^19^, or Mini Mental Status Exam (MMSE)^20^, are used inconsistently in primary care and have significant limitations; they require staff to administer and score^21^, are prone to examiner error^21–23^, and cannot capture granular data shown to be sensitive to early indicators of CI in laboratory settings, such as nuanced aspects of reaction time^24–26^. Given the extensive information clinicians must evaluate to meet the demands of clinical care and documentation, self-administered, app-based cognitive screening systems may help streamline routine cognitive screening and promote early detection in primary care settings.

MyCog, a brief, cognitive screening app self-administered on a tablet, was funded by the NIH to detect cognitive impairment in older adults during a primary care visit^27^. Adapted from the NIH Toolbox^®^ for Assessment of Neurological and Behavioral Function Cognition Battery (NIHTB-CB), MyCog consists of two cognitive tasks: (1) Dimensional Change Card Sort^28^ (MyCog DCCS), which assess executive functioning and cognitive flexibility, and (2) Picture Sequence Memory^29,30^ (MyCog PSM), which assesses episodic memory. Both measures have ample evidence of strong psychometric properties within the NIHTB^31–33^. The MyCog battery was designed to be brief (less than 7 minutes total) and easy to self-administer during the rooming process of a primary care visit. The MyCog measures have demonstrated strong evidence of reliability and construct validity, comparable to well-established, traditional cognitive assessments^34^. The two cognitive domains measured by MyCog—episodic memory (MyCog PSM) and executive function (MyCog DCCS)—align with clinical understanding of early changes associated with cognitive decline^35–37^. Episodic memory impairment is often among the earliest manifestations of Alzheimer’s pathology^35,36^, while executive dysfunction measured by MyCog DCCS captures frontal-subcortical circuit disruption common in vascular cognitive impairment and other dementias^38^. MyCog DCCS measures both speed and accuracy, which is consistent with the subtle processing inefficiencies that often precede more obvious cognitive errors in early impairment^29,30^.

MyCog has been involved in three large-scale NIH-funded grants to date (5UH3NS105562, U01NS105562, AND R01AG069762), and is currently used by three large health systems around the country, including Oak Street Health, Access Community Health, and Northwestern Medicine. The app integrates with electronic health records (including the most common EHR in the nation, Epic), and triggers customized clinical decision support to help manage flagged results. MyCog has been self-administered by over 32,000 patients in 31 primary care clinics, with promising evidence supporting its sensitivity and specificity in preliminary studies.^31^

The present study builds upon the preliminary MyCog studies via a robust clinical validation of MyCog in a combined sample of older adults with and without cognitive impairments. The goal of this study was to determine the best derived scores for differentiating between individuals with and without cognitive impairment through a systematic investigation of multiple scoring methods. We leveraged five advanced modeling techniques combining regularization, machine learning, and Bayesian approaches to identify robust predictors of cognitive impairment common across all models, which were then used to create an optimized scoring algorithm to be programmed into the MyCog application.

## Method

### Ethics Approval

All methods were approved by Northwestern University’s Institutional Review Board (STU00212972). Participants provided informed consent prior to participation. Participants in the cognitive impairment group were given an adapted form of the Brief Assessment of Capacity to Consent (UBACC) to determine eligibility to participate.^39^

### Sample

Participants were recruited from Northwestern Medical Group (NMG) Geriatrics and General Internal Medicine and Regional Medical Group (RMG) Movement Disorders and Neurodegenerative Diseases Center. The sample consisted of two groups: one with documented cognitive impairments and one without documented cognitive impairment.

Inclusion criteria for the cognitively impaired subsample required that participants: (a) were 65 years or older; (b) spoke English; (c) had an upcoming appointment (within 4 weeks of study interview) or recent appointment (within 6 months of study interview) at NMG/RMG or Neurobehavior and Memory Clinic; and (d) had a documented diagnosis in their chart of impairment classification (MCI or dementia; Table 2).

**Table 1.** Demographic information for participant samples.

|  | Cognitively impaired | Non-impaired |
| --- | --- | --- |
| Total Sample N | 65 | 80 |
| Mean Age (SD) | 79.9 (7.0) | 76.0 (5.9) |
| [Min, Max] | [67, 94] | [67, 93] |
|  | n (%) |  |
| Gender |  |  |
| Female | 36 (55.4) | 55 (68.8) |
| Male | 29 (44.6) | 25 (31.2) |
| Racial Identity |  |  |
| Asian | 2 (3.1) | 2 (2.5) |
| Black or African American | 8 (12.3) | 8 (10.0) |
| Middle Eastern or North African | 0 (0.0) | 0 (0.0) |
| Native Hawaiian or Pacific Islander | 0 (0.0) | 0 (0.0) |
| White | 53 (81.5) | 66 (82.5) |
| Other Identity | 1 (1.5) | 0 (0.0) |
| Prefer Not to Answer | 0 (0.0) | 2 (2.5) |
| Ethnic Identity |  |  |
| Hispanic | 1 (1.5) | 2 (2.5) |
| Not Hispanic | 64 (98.5) | 78 (97.5) |
| Education |  |  |
| High School Diploma or GED | 14 (21.5) | 1 (1.2) |
| Some College or 2-Year Degree | 13 (20.0) | 13 (16.2) |
| 4-Year College Degree | 18 (27.7) | 17 (21.2) |
| Graduate Degree | 20 (30.8) | 49 (61.3) |

**Table 2.** Pertinent ICD codes for cognitive deficits and impairment.

|  | Criteria (diagnosis with either ICD code) |  |
| --- | --- | --- |
|  | ICD-9 | ICD-10 |
| Dementia | 290.X, 291.0-291.9, 292.82, 294.10, 294.11, 294.20, 294.21 | F01.50, F01.51, F10.27, F10.97, F19.97, F19.27, F02.80, F02.81, F03.90, F03.913 |
| MCI | 331.83 | G31. 84 |
| Cognitive deficits | 438.0 | I69.91 |
| Other memory loss | 780.93 | R41.3 |

ICD, International Classification of Diseases; MCI, mild cognitive impairment.

Inclusionary criteria for the non-impaired subsample required that participants: (a) were 65 years or older; (b) spoke English; and (c) obtained a Montreal Cognitive Assessment (MoCA) score ≥ 26 as administered at a recent appointment (within 6 months of validation interview) at NMG/RMG, or during the validation screening procedure. For both impaired and non-impaired subsamples, exclusionary criteria included severe, uncorrected vision or hearing impairment.

### Procedure

All participants self-administered MyCog in dedicated research or clinical facilities alongside a larger battery of neuropsychological measures between the summer of 2021 and fall of 2024. For the clinical sample, diagnostic data were collected via a review of electronic medical records in the institution’s Electronic Data Warehouse.

### Measures

#### MyCog Dimensional Change Card Sort (MyCog DCCS) Task

Adapted from the NIH Toolbox Cognition Battery, the MyCog DCCS task is an executive function measure designed to assess cognitive flexibility^40^. Participants sort images by matching a center target image to one of two bottom reference images based on either color or shape. The task includes five blocks: (1) two shape practice trials, (2) five shape-only trials (pre-switch), (3) two color practice trials, (4) five color-only trials (post-switch), and (5) 30 mixed trials requiring shape or color matching in pseudo-random order. Inter-trial intervals are 1000ms for practice and single-dimension blocks, and either 300ms or 1000ms for mixed trials. Response times are measured from target onset to response, with outliers (<100ms or >10,000ms) removed. Total testing time is 2-3 minutes.

Both RT and errors are captured on the MyCog DCCS measure. The MyCog DCCS accuracy score is calculated by dividing the number of correct responses by the total number of trials to which the participant responded, with higher scores indicating better performance. The MyCog DCCS speed score is calculated as the median RT in milliseconds across the 30 mixed trials (with outliers removed), with lower speeds indicating better performances. Finally, the MyCog DCCS integrated speed-accuracy score is computed using both the rate-correct score (RCS; Equation 1)^42^ and linear integrated speed-accuracy score (LISAS; Equation 2)^43^.

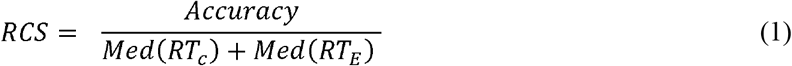

*Accuracy* refers the number of correct responses and *Med*(*RT*_*c*_) and *Med*(*RT*_*E*_) are the median RT across correct and error trials, respectively. RCS is interpreted as the number of correct responses adjusted for the total time spent. Higher RCS’s indicate better performance.

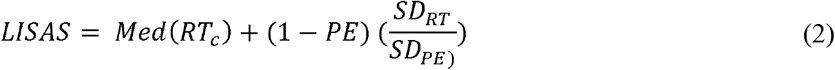

*Med*(*RT*_*c*_) is the median correct RT, is mean error proportion, *D* is the standard deviation of RT, and *D*_PE_ is the standard deviation of the error proportion. LISAS is interpreted as the median RT adjusted for error. Lower LISAS indicate better performance.

#### MyCog Picture Sequence Memory (MyCog PSM) Task

Also adapted from NIHTB-CB, the MyCog PSM task is a measure of episodic memory that requires respondents to memorize a sequence of images. The task begins with a practice trial in which 4 images are presented sequentially in pseudo-random temporal order, each accompanied by a brief audio description. All images depict easily identifiable activities with no inherent order related to the theme of “A day with a friend.” The images are presented in order with a simple audio description, then scrambled, and the patient is asked to arrange the pictures in the order they were shown. Total testing time is approximately 4 to 5 minutes. Scores considered in this study were based on the cumulative number of correct image placements (exact match) or the number of images correctly placed next to another correct image (adjacent pairs).

### Analysis

Statistical analyses were performed using R version 4.2.0. Multiple scores and their combinations were examined to determine which best contributed to predicting cognitive impairment across different trial conditions (Table 3).

**Table 3.** Description and interpretation of MyCog DCCS scores and trial conditions.

| Score | Description | Interpretation |
| --- | --- | --- |
| Proportion correct | Proportion of correct responses | Accuracy |
| Response speed | Sum of response times for correct trials (in milliseconds) | Reaction time |
| Rate-correct score (RCS) | Number of correct responses divided by mean response time (in seconds) | Speed-accuracy trade-off |
| Intraindividual standard deviation (ISD) | SD of response time for correct trials | Performance variability |
| Residualized ISD (RISD) | ISD after controlling for mean response time, practice effects, and age | Performance variability |
| Mean square successive difference (MSSD) | Average of squared differences between successive response times | Moment-to-moment variability |
| Linear integrated speed-accuracy score (LISAS) | Combined speed and accuracy measure weighted by SD | Performance error (in terms of combined speed and accuracy) |
| Difference scores (DIFF) | Differences between conditions (e.g., dominant vs non-dominant, switch vs repeat) | Performance error for harder conditions (or during switching) |
| Trial Condition |  |  |
| Total | All 30 trials, encompassing switch, repeated, dominant, and non-dominant conditions |  |
| Switch | Trials that prompt matching with a different dimension than the proceeding trial (e.g., switches from shape to color or vice versa) |  |
| Repeated | Trials that prompt matching with the same dimension than the proceeding trial |  |
| Non-Dominant | Trials that prompt matching with the color dimension (7/30 trials) |  |
| Dominant | Trials that prompt matching with the shape dimension (23/30 trials) |  |

**Table 4.** Comparison of sample performance across all study variables *M (SD), [minimum, maximum*].

| Variables / Trials | Impaired (n=65) | Non-impaired (n=80) |
| --- | --- | --- |
| MyCog Picture Sequence Memory (PSM) |  |  |
| Exact Match | 2.34 (2.27) [0.00, 9.00] | 6.24 (2.99) [2.00, 12.00] |
| Adjacent Pairs | 0.72 (1.48) [0.00, 7.00] | 3.66 (3.39) [0.00, 11.00] |
| MyCog Dimensional Change Card Sorting (DCCS) |  |  |
| Accuracy (Proportion Correct) |  |  |
| All | 0.82 (0.18) [0.33, 1.00] | 0.96 (0.08) [0.67, 1.00] |
| Switch | 0.81 (0.17) [0.31, 1.00] | 0.95 (0.09) [0.62, 1.00] |
| Non-Dom | 0.87 (0.18) [0.43, 1.00] | 0.96 (0.10) [0.43, 1.00] |
| Switch – Repeated | -0.01 (0.17) [-0.31, 0.57] | -0.01 (0.09) [-0.31, 0.41] |
| Non-Dom – Dom | 0.07 (0.27) [-0.57, 0.87] | 0.01 (0.09) [-0.31, 0.30] |
| Response Speed (Sum of Reaction Time) |  |  |
| All | 0.53 (0.26) [0.12, 1.08] | 0.92 (0.32) [0.19, 1.96] |
| Switch | 0.56 (0.28) [0.08, 1.17] | 0.90 (0.31) [0.17, 1.83] |
| Non-Dom | 0.70 (0.39) [0.07, 1.62] | 1.01 (0.35) [0.11, 2.26] |
| Switch – Repeated | 0.01 (0.24) [-0.58, 0.68] | -0.06 (0.25) [-0.93, 0.65] |
| Non-Dom – Dom | -0.19 (0.32) [-1.03, 0.61] | -0.10 (0.26) [-0.93, 0.65] |
| Rate Correct Score (RCS) |  |  |
| All | 0.53 (0.26), 0.12–1.08 | 0.92 (0.32), 0.19–1.96 |
| Switch | 0.56 (0.28), 0.08–1.17 | 0.90 (0.31), 0.17–1.83 |
| Non-Dom | 0.70 (0.39), 0.07–1.62 | 1.01 (0.35), 0.11–2.26 |
| Switch – Repeated | 0.01 (0.24), -0.58–0.68 | -0.06 (0.25), -0.93–0.65 |
| Non-Dom – Dom | -0.19 (0.32) [-1.03, 0.61] | 0.10 (0.26) [-0.93, 0.65] |
| Linear Integrated Speed-Accuracy Score (LISAS) |  |  |
| All | 1643.48 (761.80) [709.17, 4925.99] | 1029.25 (348.70) [423.33, 2035.93] |
| Switch | 1707.49 (1226.87) [689.84, 8865.86] | 1075.58 (361.05) [401.30, 2595.57] |
| Non-Dom | 1607.33 (1341.55) [602.14, 8940.32] | 1020.63 (468.14) [384.96, 4224.65] |
| Switch – Repeated | 43.12 (934.99) [-2019.70, 6427.27] | 83.70 (287.31) [-1137.29, 791.98] |
| Non-Dom – Dom | -122.88 (1056.04) [-2116.67, 6325.49] | -22.87 (401.92) [-993.23, 2411.29] |
| Intraindividual Standard Deviation (ISD) |  |  |
| All | 968.60 (517.23) [228.93, 2358.33] | 532.78 (334.39) [160.13, 1786.26] |
| Switch | 901.19 (663.52) [181.64, 2814.34] | 447.50 (275.87) [156.62, 1667.53] |
| Non-Dom | 704.23 (712.19) [85.93, 4130.55] | 341.54 (300.66) [72.82, 2439.48] |
| Residualized Intraindividual Standard Deviation (RISD) |  |  |
| All | 12.16 (6.59) [2.96, 30.64] | 6.60 (4.03) [2.08, 22.86] |
| Switch | 12.26 (9.04) [2.64, 39.08] | 6.12 (3.82) [1.63, 23.09] |
| Non-Dom | 11.52 (12.99) [1.37, 76.08] | 5.36 (4.38) [0.56, 32.97] |
| Mean Square Successive Difference (MSSD) |  |  |
| All | 2136491.90 (2302636.35)<br>[71283.85, 11673080.96] | 682646.10 (1076799.70)<br>[56814.65, 6684108.62] |
| Switch | 2533771.44 (3936278.87)<br>[72272.21, 16408903.55] | 565491.36 (952607.60)<br>[24124.17, 6679232.02] |
| Non-Dom | 2237211.36 (6790899.63)<br>[7449.36, 51055212.11] | 330911.00 (894636.97)<br>[2729.19, 7503457.81] |

#### Missing Data Assessment and Handling

Prior to model building, we assessed the extent of missing data across all variables. The initial sample included 70 patients with cognitive impairment and 81 without. Overall, missing data ranged from 0% to 2.65% (PSM exact match score) across the included variables, with an average missingness of 0.18%. Because of the low rate of missingness, the small sample, and the large number of predictive variables, we used complete cases only to conduct analyses.

#### Consensus Approach

We employed a consensus approach to identify robust predictive variables for cognitive impairment by implementing five different supervised model classes to ensure reliable variable selection and avoid model-dependent conclusions. Each model was specifically chosen for its ability to handle the relatively small sample size and correlated score types present in our dataset. We evaluated each model based on area under the receiver operating characteristic curve (ROC AUC), sensitivity, and specificity. Rather than selecting a final model based on individual performance metrics alone, we implemented a consensus strategy to identify predictors that were selected across each of the modeling approaches. This methodology increases confidence that our parsimonious final model reflects true clinical signals rather than statistical artifacts. Model stability was assessed through comprehensive agreement analysis between different approaches. We calculated pairwise agreement rates and Cohen’s kappa statistics, along with Pearson correlations between predicted probabilities across models. This multi-faceted evaluation ensured that our final variable selection was robust and reproducible across different analytical frameworks. All models used the same 5-fold cross-validation partitioning to ensure fair comparison of variable selection consistency across approaches. The following models were evaluated:

##### 1. LASSO Regression. LASSO

(Least Absolute Shrinkage and Selection Operator) was selected for its ability to perform automatic variable selection while handling multicollinearity through L1 regularization. The L1 penalty forces some coefficients to exactly zero, effectively removing irrelevant predictors from the model. Predictors underwent z-score standardization to optimize regularization performance. We employed 5-fold cross-validation using the *cv*.*glmnet* function from the *glmnet* package to select the optimal lambda (penalty strength) parameter with a default sequence of 100 logarithmically-spaced values, selecting the value that minimized cross-validated error (‘lambda.min’). Since LASSO shrinks coefficients rather than explicitly selecting variables, we applied a conservative coefficient threshold of 1×10□□ to identify practically meaningful predictors. Multicollinearity is handled by the L1 penalty, which tends to select one variable from a group of highly correlated predictors.

##### 2. Elastic Net Regression

Elastic Net combines the advantages of both LASSO and Ridge regression, making it particularly suitable for datasets with grouped variables or when the number of predictors exceeds the number of observations. Predictors underwent z-score standardization to optimize regularization performance. We used a fixed alpha value of 0.5 as the mixing parameter, balancing equally between L1 (LASSO) and L2 (Ridge) penalties, which was selected given the lack of preference between selection and shrinkage. The same cross-validation procedure as LASSO was employed for lambda selection. Elastic Net handles multicollinearity through its combined penalty structure: the L2 component stabilizes the solution when predictors are correlated, while the L1 component maintains variable selection properties.

##### 3. Random Forest

Random Forest was chosen for its ensemble approach that naturally handles correlated predictors without requiring explicit variable selection. The method uses bootstrap sampling and random feature selection to create diverse trees, reducing overfitting and handling multicollinearity through averaging across multiple decision trees. We implemented Random Forest with an adaptive number of trees (minimum 100, maximum 500, scaled by twice the training sample size) and used the default mtry parameter (equal to the square root of the total number of predictors) for feature sampling at each split. Out-of-bag error estimation provided internal validation. Multicollinearity is managed through the random sampling of both observations and features at each split, which decorrelates the individual trees and reduces the impact of highly correlated predictors.

##### 4. Bayesian Logistic Regression

Bayesian logistic regression was selected to provide additional stability in parameter estimation, particularly valuable when dealing with correlated predictors and moderate sample sizes. We used weakly informative priors via the arm package’s *bayesglm* function, which approximates the behavior of frequentist logistic regression while providing regularization through the prior distribution. The Bayesian approach naturally handles multicollinearity by incorporating prior information that shrinks coefficient estimates toward reasonable values, preventing the extreme parameter estimates that can occur with correlated predictors in maximum likelihood estimation.

##### 5. Gradient Boosting

Gradient boosting was included for its ability to capture complex non-linear relationships and interactions while maintaining interpretability through variable importance measures. We used adaptive parameter settings scaled to sample size: number of trees ranged from 50 to 100 based on training sample size, interaction depth was set to the minimum of 3 or floor(log□(number of predictors + 1)), shrinkage rate of 0.1, and bag fraction of 0.5 for stochastic gradient boosting. The method handles multicollinearity through its iterative learning process and built-in regularization: the shrinkage parameter prevents overfitting, while the random sampling (bagging) reduces the impact of correlated predictors by training each tree on different subsets of the data.

#### Final Model Development for Implementation

Finally, we selected predictors consistently identified by all five modeling approaches in over 50% of their cross-validation folds to create a consensus logistic regression algorithm. To evaluate final model performance, we used two approaches to determine optimal thresholds: 1) Youden’s J statistic, which maximizes the sum of sensitivity and specificity (Sensitivity + Specificity −1), and 2) Top-Left Point method, which identifies the threshold that minimizes the Euclidean distance to the perfect classifier point (0,1) on the ROC curve, providing a geometric optimization approach that treats sensitivity and specificity errors with equal weight. AUC was selected as the primary performance metric, with values between 0.9 and 1 considered excellent; values between 0.8 and 0.9 considered good, values between 0.7 and 0.8 considered acceptable, and values less than .7 considered unacceptable^44–46^. Additional performance metrics were calculated for each threshold method including sensitivity, specificity, accuracy, positive and negative predictive values, F1-score, and Cohen’s Kappa measure of agreement. To assess the stability of performance estimates, bootstrap confidence intervals (n=1000 iterations) were calculated with stratified sampling to maintain the original proportion of impaired to non-impaired participants in each bootstrap sample.

## Results

After applying complete case analysis to handle missing data (0.18% overall missingness), the final analytical sample consisted of 145 participants: 65 with cognitive impairment and 80 without cognitive impairment (Table 1). All five modeling approaches demonstrated strong diagnostic performance, with good to excellent ROC AUC values ranging from 0.839 to 0.876 across different model types, sensitivity ranged from 0.765 to 0.868, and specificity ranged from 0.755 to 0.866 (Table 5). Models selected between 11 (LASSO) and 22 (Gradient Boosting) variables.

**Table 5.** Comparison of diagnostic accuracy across five models.

| **Model** | **ROC AUC** | **Sensitivity** | **Specificity** | **Accuracy** | **Selected Variables (N)** |
| --- | --- | --- | --- | --- | --- |
| LASSO | 0.876 (0.050) | 0.799 (0.171) | 0.866 (0.107) | 0.828 (0.055) | 11 |
| Elastic Net | 0.865 (0.044) | 0.786 (0.166) | 0.866 (0.107) | 0.821 (0.051) | 19 |
| Random Forest | 0.840 (0.042) | 0.765 (0.176) | 0.829 (0.182) | 0.793 (0.060) | 16 |
| Bayesian Logistic | 0.853 (0.044) | 0.868 (0.170) | 0.755 (0.200) | 0.800 (0.062) | 13 |
| Gradient Boosting | 0.839 (0.055) | 0.769 (0.138) | 0.844 (0.092) | 0.814 (0.052) | 22 |

Model stability analysis revealed excellent agreement between different predictive approaches. Correlation coefficients between predicted probabilities from different models ranged from 0.789 to 0.996, indicating high consistency in risk stratification. Agreement rates between binary prediction classifications ranged from 79.3% to 100.0%, with Cohen’s kappa values between 0.578 and 1.000, suggesting substantial to excellent agreement across modeling approaches.

After testing 32 potential predictors derived from the two MyCog tasks, only two variables were consistently selected across all five modeling approaches: the MyCog PSM exact match score and the MyCog DCCS overall rate-correct score. The final consensus logistic regression model incorporating only the two universally selected predictors is shown in Equation 3:

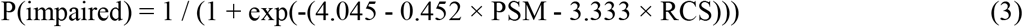

Both threshold selection methods demonstrated good discrimination (AUC = 0.890) with complementary performance characteristics and substantial agreement (Table 6). Youden’s J optimized for higher specificity (91.2%), positive predictive value (87.0%), and overall accuracy (82.8%), while the Top-Left Point method achieved higher sensitivity (78.5%) and negative predictive value (82.5%). Both approaches yielded similar F1-scores (0.790 vs. 0.785), Kappa values (0.646 vs. 0.610), and comparable accuracy (82.8% vs. 80.7%), with bootstrap confidence intervals indicating robust performance estimates for both threshold selection strategies.

**Table 6.** Final Model Performance Metrics Across Evidence-Based Thresholds with Bootstrap 95% CIs.

| **Metric** | **Youden's J** | **Top-Left Point** |
| --- | --- | --- |
| Threshold Value | .630 | .460 |
| AUC | 0.890 (0.836-0.936) | 0.890 (0.836-0.936) |
| Sensitivity | 0.723 (0.547-0.814) | 0.785 (0.678-0.877) |
| Specificity | 0.912 (0.823-0.954) | 0.825 (0.716-0.893) |
| Accuracy | 0.828 (0.738-0.869) | 0.807 (0.731-0.869) |
| PPV | 0.870 (0.767-0.930) | 0.785 (0.683-0.860) |
| NPV | 0.802 (0.705-0.859) | 0.825 (0.744-0.889) |
| F1 | 0.790 (0.647-0.857) | 0.785 (0.689-0.857) |
| Kappa | 0.646 (0.451-0.737) | 0.610 (0.449-0.726) |

## Discussion

Our findings demonstrate that MyCog (a brief, self-administered cognitive screening tool adapted from the NIH Toolbox Cognition Battery) can effectively identify cognitive impairment in older adults. All five modeling approaches demonstrated good discrimination (AUC .839 to .876), and strong agreement between the models confirmed the selected consensus variables were robust across diverse modeling approaches. The final parsimonious model incorporated just two metrics that were selected by all five models—the MyCog PSM exact match score and the MyCog DCCS overall rate correct score— and achieved good discrimination ability with an AUC of 0.89 regardless of threshold.

Depending on the threshold selection method employed, sensitivity ranged from 72.3% to 78.5% and specificity ranged from 82.5% to 91.2%, allowing clinicians to choose the optimal threshold based on their specific screening objectives (prioritizing sensitivity for case-finding versus specificity for more conservative screening). Overall accuracy was high for both approaches (80.7-82.8%), and positive predictive values ranged from 78.5% to 87.0%, indicating that most individuals flagged as impaired would indeed have cognitive impairments. Negative predictive values were also high (80.2-82.5%), suggesting reliable identification of cognitively intact individuals. Both threshold selection methods demonstrated substantial agreement between predicted classifications and actual diagnoses beyond chance prediction (Kappa = 0.610-0.646), and effective trade-off between accuracy and precision (F1-scores = 0.785-0.790).

MyCog’s diagnostic accuracy is comparable to other commonly used screening tools, which have been found to have AUC’s between .83 and .88 in meta-analyses.^47,48^ However, MyCog offers several distinct advantages over traditional tools. First, it eliminates the need for a trained examiner, reducing variability in administration and scoring, and triggers appropriate clinical decision-making support within the EHR, ensuring patients are offered proper follow-up. The self-administered nature of MyCog also helps overcome workforce constraints in busy primary care settings, where provider time is limited and specialized training in cognitive assessment may be unavailable.

Importantly, while we tested numerous derived metrics incorporating various aspects of cognitive performance (including complex intra-individual variability and condition-specific effects), our most robust model emerged from relatively straightforward scores. This simplicity enhances clinical utility and interpretability. The probabilistic formula we derived can be easily implemented within the app, enabling immediate risk stratification at the point of care. Using this formula, we evaluated two cut points that provide different sensitivity and specificity estimates. Sensitivity is commonly prioritized over specificity for initial screeners to enhance detection power at a population level,^49^ however, many of the clinicians who have worked with our team reported that specificity their priority as to not cause anxiety with false positives in an already taxed primary care setting. As such, we suggest Youden’s J cut point for a sensitivity of 72.3% and specificity of 91.2%, though MyCog could be implemented using Top Left or another evidence-based cut point based on the healthcare system’s screening needs and clinical judgement.

The ability to effectively screen for cognitive impairment in primary care settings has significant implications for addressing healthcare disparities. Early detection allows for timely intervention, management of modifiable risk factors, and appropriate care planning^50^. Moreover, as new disease-modifying treatments emerge that are only effective during mild stages of ADRD, early detection is becoming more and more critical in easily accessible primary care settings^51^.

This study has several limitations that should be considered when interpreting its results. Our sample is relatively small, and an external sample was unavailable for replication, which poses risks to generalizability. We mitigated the risk of overfitting by using a robust consensus approach to variable selection and bootstrapping confidence intervals in the final model to maximize robustness within the confines of the available sample. The sample was also predominantly White and highly educated, particularly in the non-impaired clinical group, which may not reflect the demographic diversity seen in many primary care clinics. However, MyCog’s current use in with over 32,000 patients across three large and diverse healthcare systems supports its feasibility for patients across demographic groups, and validations in specific subpopulations are warranted. It is also important to note that the cross-sectional design cannot address the predictive validity of MyCog for detecting future development of cognitive decline or conversion to dementia. Longitudinal studies are needed to determine whether MyCog can identify individuals at risk for progression before symptoms impact daily functioning.

MyCog demonstrates strong diagnostic accuracy for detecting cognitive impairment in older adults through a parsimonious model focusing on episodic memory and executive function. By addressing key implementation barriers in primary care settings—requiring only minutes to self-administer during the rooming process—MyCog has potential to increase screening rates and facilitate earlier detection of cognitive impairment. As a validated tool that integrates with electronic health records and triggers appropriate clinical decision support, MyCog represents a practical solution to improve cognitive health monitoring in primary care.

## Data Availability

All data produced in the present study are available upon reasonable request to the authors

## Acknowledgments

This work was funded by a grant from the National Institute of Health’s National Institute on Aging (R01AG069762). The funding agency played no role in the study design, collection of data, analysis, or interpretation of data.

